# Association of an Automated Breast Arterial Calcification Score with Cardiovascular Outcomes and Mortality

**DOI:** 10.1101/2023.09.29.23296371

**Authors:** Quan M. Bui, Tara Shrout Allen, Richard Mantey, Gregory M. Petersen, Junhao Wang, Nitesh Nerlekar, Mohammad Eghtedari, Lori B. Daniels

## Abstract

**Background:** Breast arterial calcification (BAC), visible on mammograms, has emerged as a biomarker of cardiovascular disease (CVD) in women. Barriers to clinical implementation of BAC include limited studies with clinical outcomes and lack of quantification tools.

**Methods:** This single-center, retrospective study included women with a screening digital mammogram from 2008-2016. BAC was quantified using an automated, artificial intelligence (AI)-generated Bradley score, as a binary (Bradley score ≥5) and continuous variable. Clinical outcomes were determined via electronic medical records. Regression analyses were used to evaluate the association between BAC and outcomes of mortality and a composite of acute myocardial infarction, heart failure, stroke, and mortality. Models were adjusted for age, race, diabetes, smoking status, blood pressure, cholesterol, and history of CVD and chronic kidney disease.

**Results:** A total of 18,092 women were included with a mean age 56.8±11.0 years. Prevalence of comorbidities included diabetes (13%), hypertension (36%), hyperlipidemia (40%) and smoking (5%). BAC was present in 4,223 (23%). Over a median follow-up of 6 years, death occurred in 7.8% women with and 2.3% women without BAC. The composite outcome occurred in 12.4% of women with and 4.3% of women without BAC. Compared to those without, women with BAC had adjusted hazard ratios (aHR) of 1.49 (95% CI 1.33-1.67) for mortality and 1.56 (1.41-1.72) for the composite endpoint, after accounting for traditional risk factors. With a continuous BAC score, each 10-point increase was associated with higher risk of mortality (1.08 [1.06-1.11]) and the composite endpoint (1.08 [1.06-1.10]). BAC was especially predictive of future events among younger women.

**Conclusion:** BAC is significantly and independently associated with mortality and incident CVD, especially among younger women. Measuring BAC using an AI algorithm is feasible and clinically relevant. Further studies are needed to confirm these findings and to evaluate whether interventions guided by BAC improve outcomes.

**CLINICAL PERSPECTIVE:** *What is new?:*

- Breast arterial calcification (BAC) on mammograms can be reliably quantified using a novel software based on an artificial intelligence (AI) algorithm.
- BAC is independently associated with an increased risk of all-cause mortality and cardiovascular outcomes. These associations held true when looking at BAC as presence, score quartile, and as a continuous value as well as after accounting for traditional cardiovascular risk factors.
- In stratification analysis, BAC was most predictive of all-cause mortality and cardiovascular outcomes among younger women (age 40-59 years), but still independently predictive in women aged 60-74 years.

*What are the clinical implications?:*

- Our data provide support for the inclusion of BAC findings on mammogram reports.
- Automated quantification tools and reporting methods of BAC will be critical to engagement of radiologists and implementation of reporting.
- While additional studies are needed to determine the appropriate clinical response, the presence of BAC should at the minimum stimulate patient-provider conversations on lifestyle changes to mitigate cardiovascular risk, especially among younger women.

## INTRODUCTION

Cardiovascular disease (CVD) remains the leading cause of death in women despite significant advances in cardiovascular diagnostics and treatments.^1^ Delays in diagnosis and treatment, as well as under-treatment, contribute to morbidity and mortality.^2^ This problem is further exacerbated by under-representation of women in cardiovascular clinical trials and lack of sex-specific CVD screening tools.^3^ Methods to broadly screen women that are efficient and effective are sorely needed.

Breast arterial calcification (BAC), an incidental finding on mammograms, has emerged as a sex-specific biomarker for atherosclerotic cardiovascular disease (ASCVD) that offers the potential for personalized risk stratification.^4^ Gleaning information from an imaging study beyond its original intent is not new; analogous to BAC on mammography is coronary artery calcifications seen on chest computed tomography obtained for non-cardiac purposes.^5^ BAC has tremendous appeal because it is non-invasive, comes at no additional cost or radiation, and the majority of women over the age of 40 years already undergo annual screening mammography for breast cancer.^6^

Multiple studies have found significant associations between the presence of BAC and prevalent CVD.^4^ It is postulated that BAC represents lifetime exposure to risk factors related to arterial stiffening, which increases the risk of CVD through both coronary and non-coronary mechanisms (i.e. heart failure and stroke).^7^ However, routine clinical use of BAC has not been adopted due to a lack of outcomes studies as well as technological challenges in measuring and reporting BAC.^4^ The purpose of this study was to evaluate the association of BAC with CVD risk factors and hard clinical outcomes as well as to validate an automated, artificial intelligence (AI) algorithm for BAC quantification.

## METHODS

### Study Population

This single-center retrospective study included women between the ages of 40 and 90 years who underwent screening digital mammography between 2008 to 2016 at University of California (UC) San Diego Health. For each subject, only the index mammogram was analyzed. All protocols were approved by the Institutional Review Board of UC San Diego Health (IRB #170154).

### Evaluation of BAC

BAC was quantified using a validated, proprietary investigational software (cmAngio^®^, CureMetrix) based on a deep neural, AI network and previously trained using over 21,000 2D full-field digital mammograms (FFDM) obtained from multiple sites in Australia, Brazil, and the United States (not including UC San Diego Health).^8^ For the present study, four FFDM images from each participant were used. The software cmAngio™ assesses screening mammography images and feeds them through the deep learning model to identify regions of interest within the breast. These regions correspond to areas that the algorithm suspects to have a high probability of BAC. From these identified regions, local and global imaging features such as density, contrast, and other physical dimensions are combined to determine the severity of BAC. This process is applied to each of the four standard screening mammography images. Following these calculations, each image is assigned a score between 0-100 corresponding to the severity of the BAC finding(s), with 0 representing no BAC and 100 representing a level of BAC within the highest percentile. To balance the algorithm’s false positive and false negative rate, all image-level scores less than 5 are floored to 0. The patient level score (or Bradley Score) is the mean of the threshold image-level scores across all 4 views. In this study, BAC was evaluated both as a binary (presence vs absence), and as a continuous variable (Bradley Score 0-100), as well as by quartiles (1^st^ to 4^th^). BAC presence was defined as a mean Bradley Score ≥ 5. Scores were distributed by severity into the following quartiles: 1^st^ Quartile [Score 1-25], 2^nd^ Quartile [Score 26-50], 3^rd^ Quartile [Score 51-75], and 4^th^ Quartile [Score 76-100].

### Clinical Data and Outcomes

The primary outcome was all-cause mortality, which is thereafter referenced as mortality. Secondary outcomes included acute myocardial infarction (MI), heart failure, stroke, and a cardiovascular composite outcome (MI, heart failure, stroke, and mortality). Those with baseline MI, heart failure, and/or stroke were excluded from the relevant outcome and composite analyses. Outcomes were collected via electronic medical records (EMR) using International Classification of Disease ICD-10 diagnoses until death or the censoring date of December 31^st^, 2020. Additional clinical data including baseline characteristics, medical history, vital signs and labs were collected using the EMR.

### Analyses and Statistical Methods

Continuous variables were reported either as mean with standard deviation or as median with interquartile range as appropriate based on normality of distribution assessed by Shapiro-Wilk test. Categorical variables were expressed as counts with percentages. Variables were compared using the unpaired Student t-test, Mann-Whitney test, and Fisher exact test, as appropriate. Kaplan-Meier curves were plotted, and Cox Proportional Hazards regression analyses were used to determine associations between BAC (as a binary and continuous variable) and clinical outcomes, sequentially controlling for age, race/ethnicity, smoking status, systolic blood pressure (SBP), diastolic blood pressure (DBP), total cholesterol, low density lipoprotein (LDL) cholesterol, diabetes mellitus, and history of CVD or CKD, as measured at the time of FFDM.

Age was continuous and measured in years. Smoking status was categorical and defined as current, former, never, or unknown. SBP and DBP were continuous and measured in mmHg. Total cholesterol and LDL cholesterol were continuous and measured in mg/dL. Diabetes mellitus and CKD were defined as per the relevant ICD code. CVD was defined as an ICD code for any of the following: atherosclerotic cardiovascular disease (ASCVD), MI, acute ischemic heart disease, cerebrovascular disease, coronary artery disease (CAD), heart failure, or stroke. For those without covariate data from the time of FFDM, imputation was performed to account for these missing data. Data were imputed by training a nearest neighbor multiple-imputation model in Python to predict missing variables using the 10 nearest neighbors based on the collected diagnosis codes, age, ethnicity, smoking status, blood pressure (systolic and diastolic), and cholesterol (total and LDL).

Forest plots were created to assess the association between BAC and outcomes, stratified by subgroups of baseline characteristics. Tails represent 95% confidence intervals [CIs]. All reported p-values were two-sided with a value of <0.05 considered statistically significant. Statistical analyses and figures were completed using Python 3.11.5 with packages including Pandas 2.1.0 and SciPy 1.11.2.

## RESULTS

### Study Population

A total of 21,438 screening mammograms were obtained between 2008 and 2016. Of these, 1,546 mammograms were excluded for age and 1,800 mammograms were excluded for not being the index study. Therefore, 18,092 women with index mammograms were included in the study (**Figure 1**).

**Figure 1.**
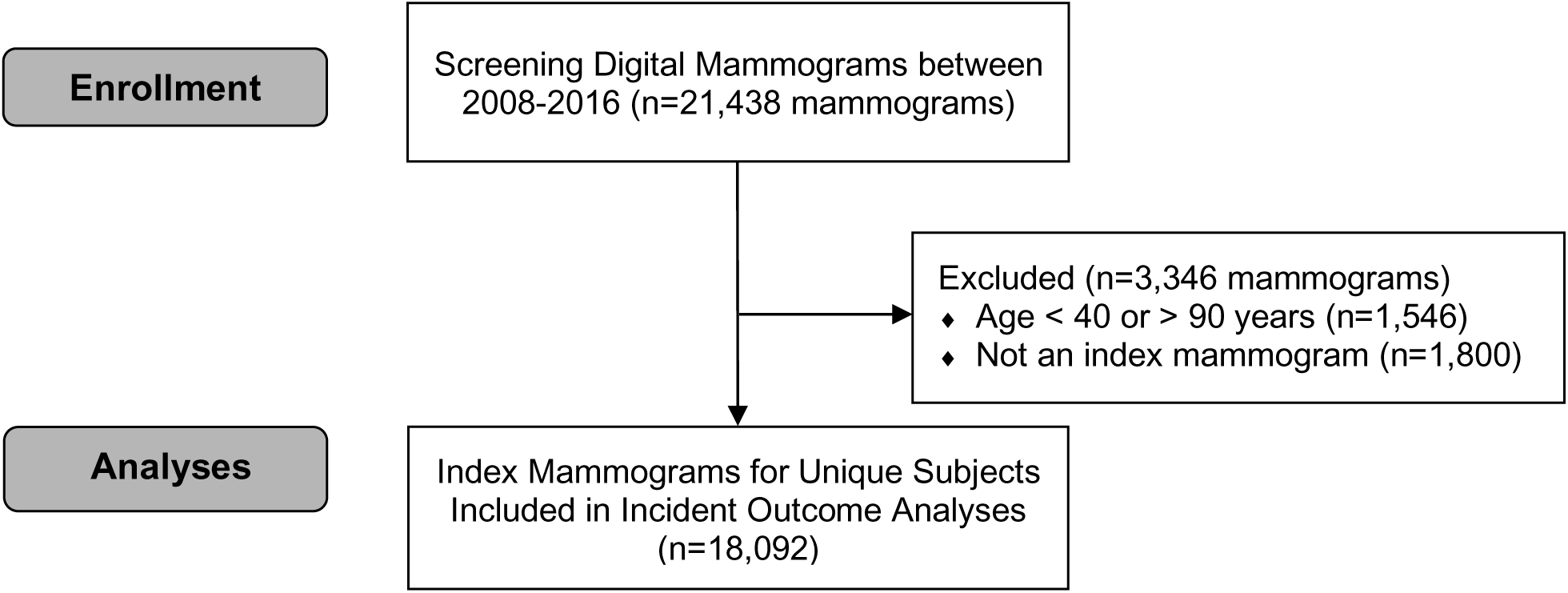
Participant Flow Diagram. After exclusions for age and non-index mammograms, there were 18,092 unique women with index mammograms included in this study.

Among the 18,092 women included, the mean age was 56.8 ±11.0 years; prevalent CVD risk factors included diabetes (13%), hypertension (36%), and hyperlipidemia (40%) (**Table 1**). BAC was present in 4,223 (23%). BAC was more prevalent among women who were older, Black or Hispanic, diabetic, hypertensive, with a history of CVD or CKD, and among those taking cardiac medications. BAC was less prevalent in current smokers. Among those with BAC, the median Bradley score was 15 (IQR 4, 50). Scores were distributed by severity into the following quartiles: 1^st^ Quartile [Score 1-25], n=2552 (60.4%); 2^nd^ Quartile [Score 26-50], n=643 (15.2%); 3^rd^ Quartile [Score 51-75], n=509 (12.1%); and 4^th^ Quartile [Score 76-100], n=519 (12.3%). Those with a higher Bradley Score were more likely to be older, have diabetes, hypertension, hyperlipidemia, history of CVD and CKD, and take statin and antihypertensive medications. (**Supplementary Table 1**).

**Table 1.** Baseline Participant Characteristics by Presence of Breast Arterial Calcification.

| Baseline Characteristics | Total<br>(n=18,092) | BAC Present<br>(n=4,223; 23%) | BAC Absent<br>(n=13,869; 77%) | *p* |
| --- | --- | --- | --- | --- |
| Age, mean years (SD) | 56.8 (11.4) | 65.2 (11.6) | 54.2 (10.0) | <0.001 |
| **Race/Ethnicity** |  |  |  |  |
| Caucasian, No. (%) | 11,319 (62.6) | 2,617 (62.0) | 8,702 (62.7) | 0.38 |
| Black/African American, No. (%) | 907 (5.0) | 241 (5.7) | 666 (4.8) | 0.02 |
| Hispanic/Latino, No. (%) | 1,694 (9.4) | 455 (10.8) | 1,239 (8.9) | <0.001 |
| Asian/Pacific Islander, No. (%) | 2,321 (12.8) | 496 (11.8) | 1,825 (13.2) | 0.02 |
| Other, No. (%) | 1,851 (10.2) | 414 (9.8) | 1,437 (10.4) | 0.31 |
| Diabetes, No. (%) | 2,267 (12.5) | 730 (17.3) | 1,537 (11.1) | <0.001 |
| Hypertension, No. (%) | 6,529 (36.1) | 2,179 (51.6) | 4,350 (31.4) | <0.001 |
| Hyperlipidemia, No. (%) | 7,256 (40.1) | 2,071 (49.0) | 5,185 (37.4) | <0.001 |
| History of CVD, No. (%) | 874 (4.8) | 424 (10.0) | 450 (3.2) | <0.001 |
| History of CKD, No. (%) | 802 (4.4) | 358 (8.48) | 444 (3.2) | <0.001 |
| Current smoking, No. (%) | 834 (4.6) | 134 (3.17) | 700 (5.1) | <0.001 |
| Never smokers, No. (%) | 9,245 (51.1) | 2,046 (48.5) | 7,199 (52.6) | <0.001 |
| SBP, mmHg (IQR) | 123 (21) | 128 (20) | 122 (20) | <0.001 |
| Total cholesterol, mg/dL (IQR) | 198 (52) | 194 (53) | 199 (51) | <0.001 |
| Statin use, No. (%) | 3,947 (21.8) | 1,430 (33.9) | 2,517 (18.1) | <0.001 |
| Antihypertensive use, No. (%) | 3,498 (19.3) | 1,313 (31.1) | 2,185 (15.8) | <0.001 |
| BAC, breast arterial calcification; SD, standard deviation; No, number; IQR, interquartile range; CKD, chronic kidney disease; CVD, cardiovascular disease; SBP, systolic blood pressure. |  |  |  |  |

### Clinical Outcomes

Over a median follow-up for mortality of 6 years [range, 6 months – 12 years], there were 329 deaths in those with BAC (7.8%) and 313 deaths in those without BAC (2.3%) (**Table 2).** Over a median follow-up for the CVD composite outcome of 4.4 years [range, 6 months – 12 years], there were 500 events (12.4%) in those with BAC and 582 events in those without BAC (4.3%). Kaplan-Meier Plots for mortality and the composite outcome are shown in **Figure 2**, which demonstrate a significantly increased risk of outcomes in those with BAC (p<0.001 for each). Kaplan-Meier Plots for individual MI, Stroke, and HF outcomes are shown in **Supplementary Figure 1**, which also demonstrate significantly increased risk in those with BAC (p<0.001 for each outcome).

**Figure 2.**
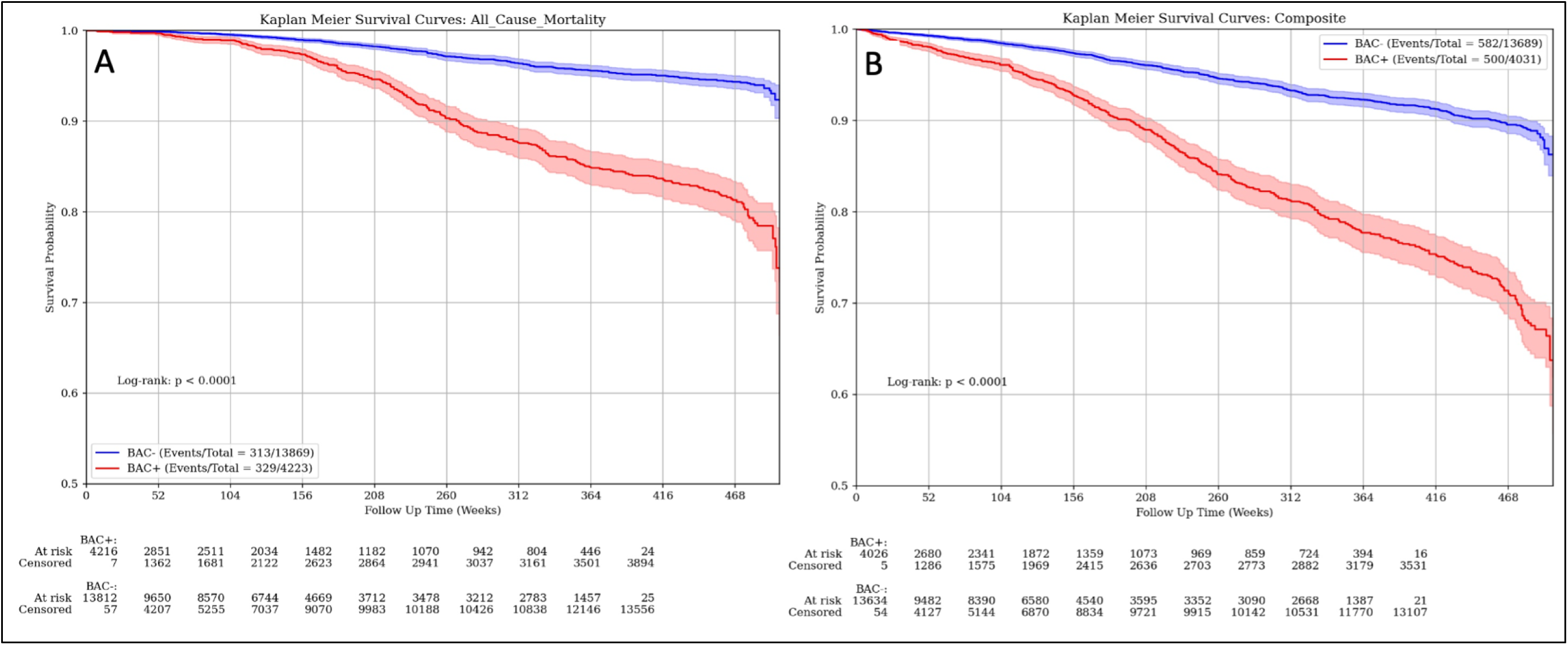
Kaplan-Meier Plots for Mortality and Composite Outcome by Breast Arterial Calcification Presence. Risk for (**A**) mortality, and (**B**) the cardiovascular composite outcome significantly varied by the presence of breast arterial calcification (p<0.001 for each). The composite outcome included acute myocardial infarction, heart failure, stroke, and mortality. BAC indicates breast arterial calcification; BAC+, presence of BAC and BAC-, absence of BAC.

**Table 2.** Clinical Outcomes by Breast Arterial Calcification Presence.

| Incident Clinical Outcomes | Total<br>(n=18,092) |  | BAC Present<br>(n=4,223) |  | BAC Absent<br>(n=13,869) |  | *p* |
| --- | --- | --- | --- | --- | --- | --- | --- |
| Myocardial infarction, No. (%) | 18,051 | 83 (0.5%) | 4,204 | 36 (0.9%) | 13,847 | 47 (0.3%) | <0.001 |
| Heart failure, No. (%) | 17,911 | 298 (1.7%) | 4,119 | 154 (3.7%) | 13,792 | 144 (1.0%) | <0.001 |
| Stroke, No. (%) | 17,914 | 259 (1.5%) | 4,138 | 110 (2.7%) | 13,776 | 149 (1.1%) | <0.001 |
| Mortality, No. (%) | 18,092 | 642 (3.6%) | 4,223 | 329 (7.8%) | 13,869 | 313 (2.3%) | <0.001 |
| Composite Outcome*, No. (%) | 17,720 | 1,082 (6.1%) | 4,031 | 500 (12.4%) | 13,689 | 582 (4.3%) | <0.001 |
| BAC, breast arterial calcification; No, number. *The cardiovascular composite outcome included acute myocardial infarction, heart failure, stroke, and mortality. |  |  |  |  |  |  |  |

On multivariable analysis, women with BAC had a higher risk of mortality (aHR 1.49 [95% CI 1.33-1.68]) and the composite outcome (aHR 1.57 [95% CI 1.42-1.74]), compared to those without BAC, in fully adjusted models (**Table 3**). With BAC quantified by quartiles, there was a graded increased risk for mortality and the composite outcome with increasing quartile, even after adjustment for cardiovascular risk factors (**Table 4**). When analyzed as a continuous Bradley score, each 10-point increase in BAC was significantly and independently associated with higher risk for adverse clinical outcomes: mortality (aHR 1.08 [95% CI 1.06, 1.11], p < 0.001) and composite outcome (aHR 1.08 [95% CI 1.06, 1.10], p < 0.001) (**Table 4)**. Similar associations with BAC were seen for heart failure and stroke, though results for myocardial infarction (n=83 events) did not reach statistical significance. (**Supplementary Tables 2 and 3**).

**Table 3.**
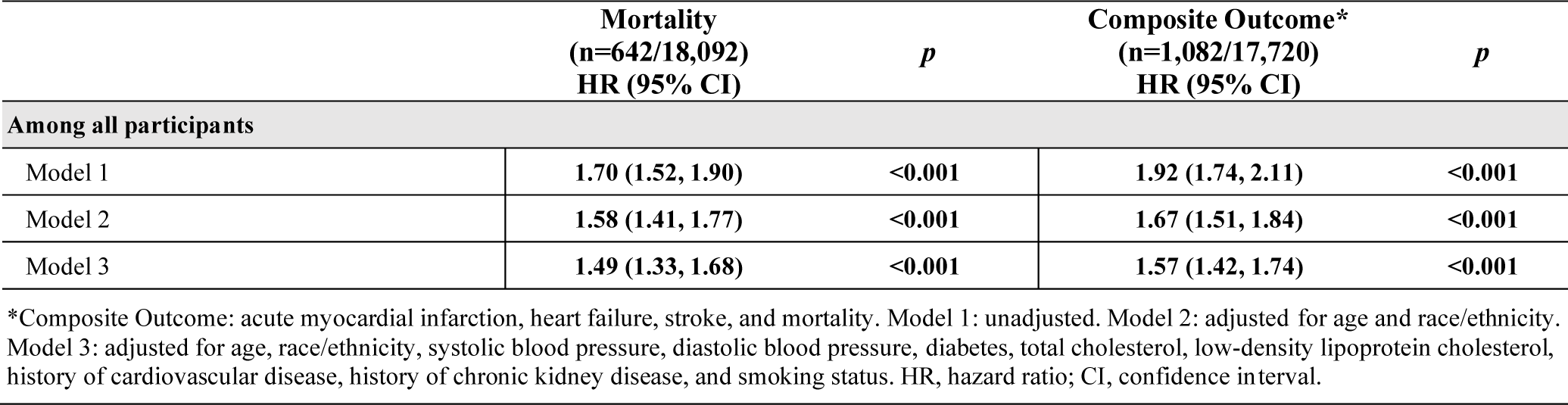
Association of Breast Arterial Calcification and Mortality and Composite Outcome.

|  | Mortality<br>(n=642/18,092)<br>HR (95% CI) | *p* | Composite Outcome*<br>(n=1,082/17,720)<br>HR (95% CI) | *p* |
| --- | --- | --- | --- | --- |
| **Among all participants** |  |  |  |  |
| Model 1 | 1.70 (1.52, 1.90) | <0.001 | 1.92 (1.74, 2.11) | <0.001 |
| Model 2 | 1.58 (1.41, 1.77) | <0.001 | 1.67 (1.51, 1.84) | <0.001 |
| Model 3 | 1.49 (1.33, 1.68) | <0.001 | 1.57 (1.42, 1.74) | <0.001 |

\*Composite Outcome: acute myocardial infarction, heart failure, stroke, and mortality. Model 1: unadjusted. Model 2: adjusted for age and race/ethnicity. Model 3: adjusted for age, race/ethnicity, systolic blood pressure, diastolic blood pressure, diabetes, total cholesterol, low-density lipoprotein cholesterol, history of cardiovascular disease, history of chronic kidney disease, and smoking status. HR, hazard ratio; CI, confidence interval.

**Table 4.** Association of Bradley Score Quartiles and Mortality and Composite Outcome.

|  | Mortality<br>(n=642/18,092)<br>HR (95% CI) | *p* | Composite Outcome*<br>(n=1,082/17,720)<br>HR (95% CI) | *p* |
| --- | --- | --- | --- | --- |
| BAC negative, n=13,869 | *Referent* | -- | *Referent* | -- |
| Per 10-point Bradley Score Increase | 1.08 (1.06, 1.11) | <0.001 | 1.08 (1.06, 1.10) | <0.001 |
| 1<sup>st</sup> Quartile [Score 1-25], n=2552 | 1.22 (1.06, 1.41) | 0.006 | 1.26 (1.11, 1.43) | <0.001 |
| 2<sup>nd</sup> Quartile [Score 26-50], n=643 | 1.44 (1.13, 1.85) | 0.004 | 1.74 (1.42, 2.13) | <0.001 |
| 3<sup>rd</sup> Quartile [Score 51-75], n=509 | 1.69 (1.33, 2.14) | <0.001 | 1.83 (1.49, 2.25) | <0.001 |
| 4<sup>th</sup> Quartile [Score 76-100], n=519 | 2.27 (1.81, 2.85) | <0.001 | 2.30 (1.88, 2.82) | <0.001 |

\*Composite Outcome: acute myocardial infarction, heart failure, stroke, and mortality. All data from the multivariable-adjusted model (Model 3), which adjusted for age, race/ethnicity, systolic blood pressure, diastolic blood pressure, diabetes, total cholesterol, low-density lipoprotein cholesterol, history of cardiovascular disease, history of chronic kidney disease, and smoking status. HR, hazard ratio; CI, confidence interval.

### Breast Arterial Calcification and Clinical Outcomes Among Subgroups

BAC prediction for mortality and the composite cardiovascular outcome significantly varied by age, SBP, total cholesterol, LDL cholesterol, smoking, and diabetes (p-interaction terms <0.001 for each). Additionally, prediction significantly varied by history of CVD for mortality (p-interaction term <0.001) and the composite outcome (p-interaction term 0.009). While prediction also significantly varied by history of CKD for mortality (p-interaction term 0.004), it did not for the composite outcome (p-interaction term 0.16). Kaplan-Meier Plots for mortality and the composite outcome stratified by age groups (**Figure 3**) demonstrate a significant separation of curves for women aged 40-59 and 60-74 years of age (p<0.001), but not for those aged 75-90 years (morality, p=0.10; composite, p=0.05). Forest plots demonstrating adjusted hazard ratios (aHR) for outcomes by stratification of baseline characteristics are shown in **Figure 4**. When stratified by age groups, and after accounting for traditional risk factors, those in the youngest age group of 40-59 years had the highest residual risk associated with BAC (mortality: aHR 1.51, 95% CI 1.22-1.87; composite outcome: aHR 1.52, 95% CI 1.25-1.85). There remained significantly increased risk associated with BAC beyond traditional risk factors for women aged 60-74 years (mortality: aHR 1.26, 95% CI 1.06-1.50; composite outcome: aHR 1.36, 95% CI 1.18-1.58), but not among those aged 75 to 90 years (mortality: aHR 1.19, 95% CI 0.91-1.54; composite outcome: aHR 1.23, 95% CI 0.98-1.55). When stratified by several other baseline characteristics, including systolic blood pressure and diabetes, the association between BAC and future cardiovascular events remained robust, even after accounting for traditional risk factors (**Figure 4**).

**Figure 3.**
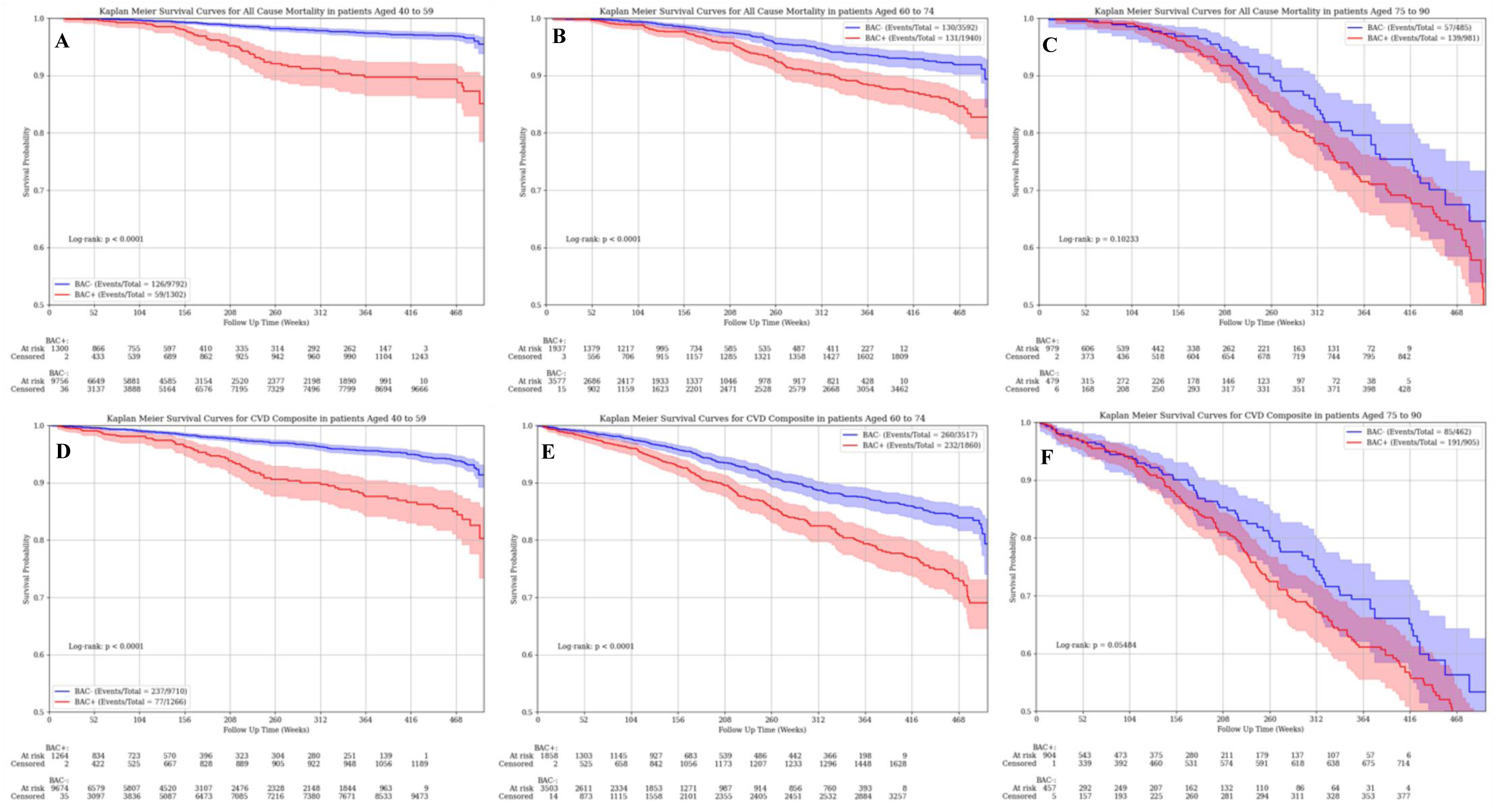
Association of Breast Arterial Calcification and Mortality and Cardiovascular Composite Outcome Stratified by Age Groups. Risk for mortality (**A-C**) and the cardiovascular composite outcome (**D-F**) by breast arterial calcification (BAC) presence/absence. Risk for both outcomes significantly varied by BAC status among women aged 40-59 years (**A, D**) and those aged 60-74 years (**B, E**) (p<0.001 for each); however, among women aged 75-90 years (**C, F**), there was no significant difference in risk for either outcome by BAC status. The composite outcome included acute myocardial infarction, heart failure, stroke, and mortality. BAC indicates breast arterial calcification; BAC+, presence of BAC and BAC-, absence of BAC.

**Figure 4.**
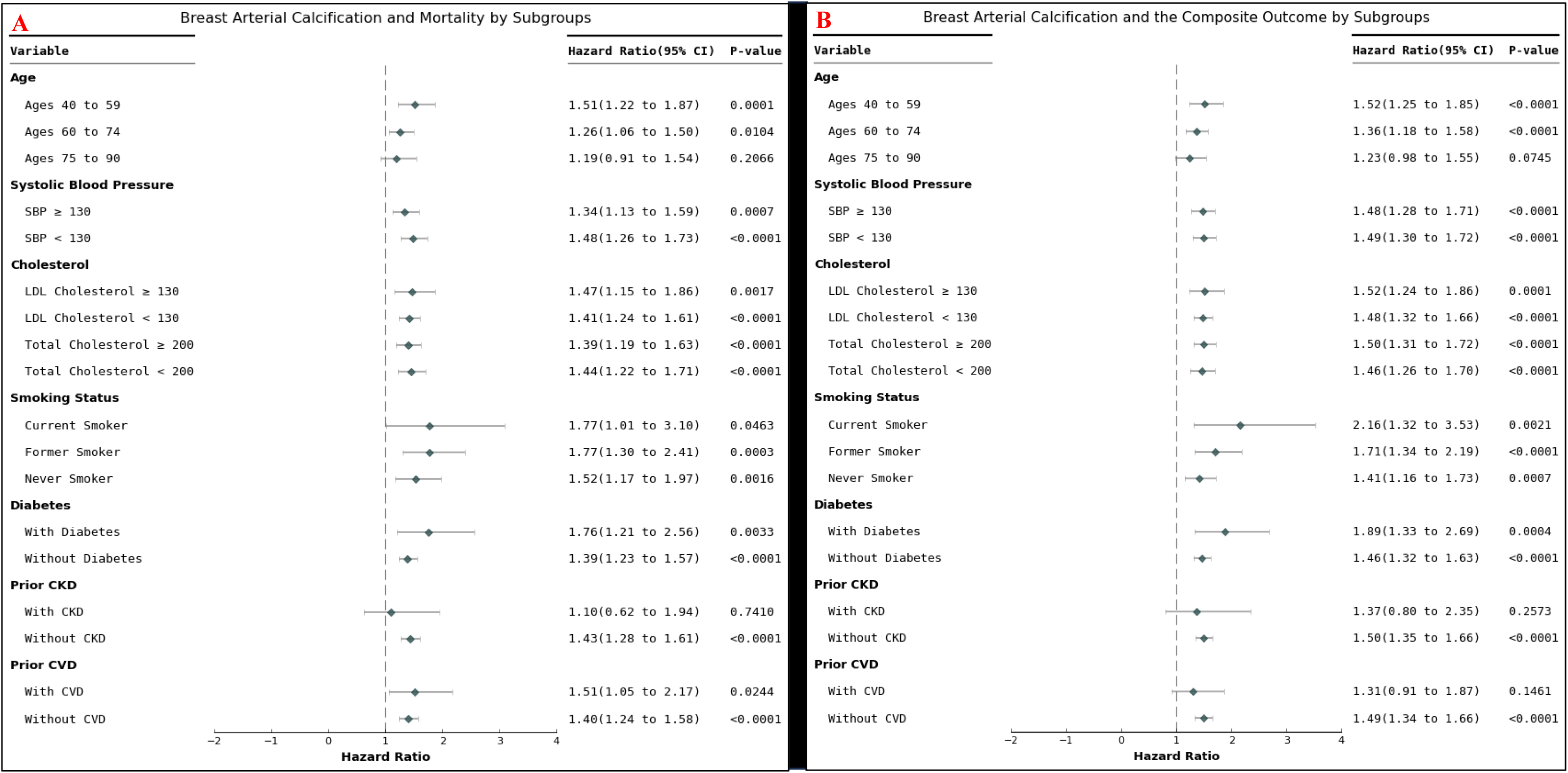
Association of Breast Arterial Calcification and Mortality and Cardiovascular Composite Stratified by Baseline Characteristics. Adjusted hazard ratios (HRs) for (**A**) mortality and **(B)** the cardiovascular composite outcome by breast arterial calcification (BAC) presence versus absence are presented. The composite outcome included acute myocardial infarction, heart failure, stroke, and mortality. BAC indicates breast arterial calcification; BAC+, presence of BAC and BAC-, absence of BAC. HRs presented were adjusted for age, race/ethnicity, systolic blood pressure, diastolic blood pressure, diabetes, total cholesterol, low-density lipoprotein cholesterol, smoking status, and history of cardiovascular disease, history of chronic kidney disease.

## DISCUSSION

In this large, retrospective study, the presence and quantity of BAC were significantly associated with all-cause mortality and the CVD composite outcome, even after adjusting for established cardiovascular risk factors. The prevalence of BAC was 23%, which constitutes a substantial proportion of women (mean age of 56.8 years) undergoing routine screening mammography. To our knowledge, this is the first study to demonstrate a significant, independent relationship between a quantitative BAC score and all-cause mortality as well as a CVD composite outcome. For example, each 10-point increase as well as sequential quartiles of the Bradley score were both significantly associated with higher risk of mortality, highlighting the potential utility of BAC quantification for personalized risk assessment.

Prior studies have only evaluated the association of BAC using a binary or a semiquantitative approach (such as absence, slight, moderate, and severe intensity) with CVD outcomes ^4,9^. In the present study, BAC was quantified using an automated method driven by a trained deep neural AI network, recently validated with high diagnostic performance.^8^ Other machine learning techniques have been developed for BAC quantification, including a densitometry method, and have been validated prospectively.^10^ Such studies have assessed methods of BAC quantification, though await association with clinical outcomes.^11–13^ The findings in our study support the efficacy of assessing both BAC presence and quantitative BAC Bradley score to improve risk assessment for mortality and CVD outcomes in women undergoing screening mammography. With the advent of AI in medical imaging, automated, quantitative BAC assessment may facilitate seamless integration into clinical workflow and allow personalized risk assessment.

Importantly, this large study also demonstrated the association of BAC with CVD outcomes and mortality even among subgroups not already known to be “high risk”, including younger women, non-smokers, and those without diabetes, high blood pressure, hyperlipidemia, CKD, or known CVD. We found that BAC was most predictive of future events among those in the youngest age group of 40-59 years, though BAC was also an independent predictor among women ages 60-74. Our results are concordant with those of Minssen et. al., who found that the diagnostic accuracy (∼84%) for BAC with CAC was the highest in patients under the age of 60 years.^14^ These findings are important, since they suggest that early risk-stratification with BAC in younger women may help identify new candidates for lifestyle modification and preventative therapies, and may ultimately help improve their outcomes. Indeed, results from this study and others suggest that BAC may develop at an earlier age than other traditional cardiovascular risk factors, and thus could serve as an early sign of underlying ASCVD risk.^15^ Similarly, the independent association of BAC with outcomes even among women without various traditional comorbidities may provide opportunities to identify risk and prevent disease in a previously unrecognized subgroup of women.

Even with engagement from cardiologists and patients, the success of BAC implementation hinges on buy-in and education of the radiology community. A survey of the members of the Society of Breast Imaging found that 85% were aware of the association of BAC with CVD, but only 15% routinely included BAC data on mammogram reports.^16^ One of the major barriers to universal BAC reporting is the lack of radiology society guidelines on reporting and appropriate use of BAC.^5,16^ Automated quantification and reporting methods for BAC will be critical to ensure that the current radiology workflow is not compromised.^7^ Therefore, it will be important for cardiologists to advise and collaborate with the breast imaging community to develop clear BAC reporting guidelines and apply automated quantification tools into clinical workflow.

If the development and implementation of BAC can follow a similar pathway as CAC, BAC may someday be used to improve CVD risk stratification beyond current tools such as the Pooled Cohort Equation (PCE), the ASCVD Risk Score, and the Framingham Risk Score (FRS). Reclassification of risk will help identify those who will benefit from more aggressive lifestyle modifications and medical therapy (i.e., statins, antihypertensives). Recently, the Multiethnic Study of Breast Arterial Calcium Gradation and Cardiovascular Disease (MINERVA) demonstrated that presence of BAC conferred additional risk at every category (i.e. low, medium and high risk) of the PCE.^10^ While our study did not include current CVD risk stratification tools, we found that BAC was independently associated with mortality and various CVD outcomes. Future studies should assess whether BAC could be used to help reclassify intermediate ASCVD risk patients to guide initiation of statins, similar to CAC as suggested in the 2018 ACC/AHA Cholesterol Guidelines.^17^ Ultimately, BAC may offer important risk stratification information without additional cost or radiation.^18^

Our study has a few limitations to note. First, the retrospective nature of the study does not prove causality. Although attempts at reducing confounding using multivariable models were used, residual risk remains. Clinical data including outcomes relied on the use of ICD-10 codes from an EMR data extraction which introduces the possibility of misclassification. Also, mortality information only included all-cause mortality; we did not have data on cause-specific mortality including CVD death. Additionally, most subjects in this study identified as white, making results most applicable to this population. Future research should focus on the implications of BAC across more diverse populations to further support external validity of this potential screening tool and to identify additional areas of opportunity.

## CONCLUSION

In this large, retrospective study, both BAC presence and quantity are significantly and independently associated with mortality and a CVD composite outcome. BAC appears to be especially predictive of CVD risk among younger women. Reporting of BAC was feasible and reliable using an automated AI algorithm, which could facilitate reporting uptake within the radiology community. Further studies are needed to determine the appropriate clinical response to BAC, and whether such a response can improve CVD outcomes in women.

## Data Availability

The data that support the findings of this study are available from the authors and CureMetrix. Restrictions apply to the availability of these proprietary data, which were used under license for this study. Data are available from the authors with the permission of CureMetrix and subject to individualized data use agreements.

## ABBREVIATIONS

BAC: breast arterial calcification
CVD: cardiovascular disease
AI: artificial intelligence
aHR: adjusted hazard ratios
ASCVD: atherosclerotic cardiovascular disease
FFDM: full-field digital mammograms
MI: acute myocardial infarction
HF: heart failure
SBP: systolic blood pressure
DBP: diastolic blood pressure
LDL: low-density lipoprotein cholesterol
CKD: chronic kidney disease
EMR: electronic medical records
UCSD: University of California, San Diego

---

**Supplementary figure S1.**
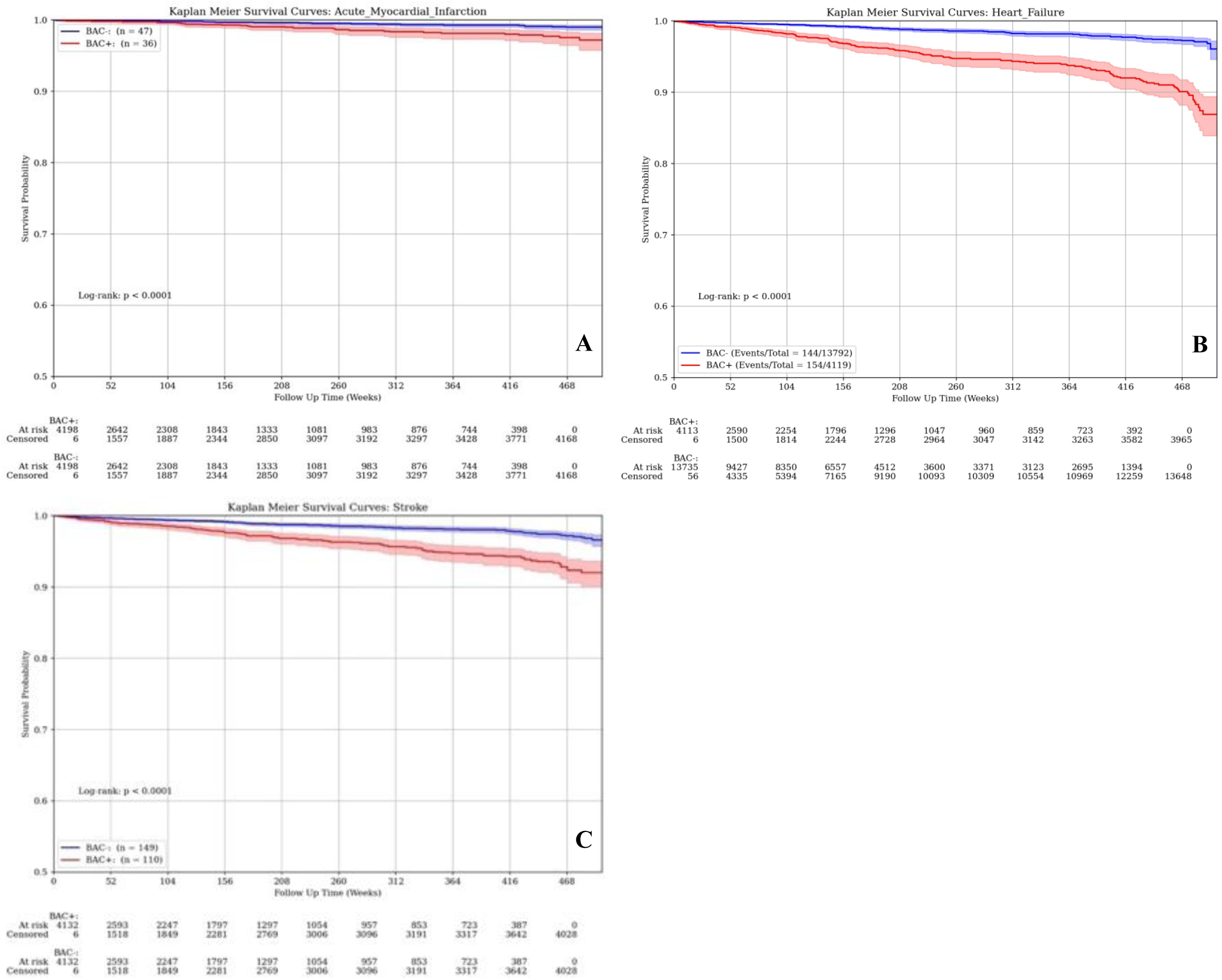
Kaplan-Meier Plots for Acute Myocardial Infarction, Heart Failure, and Stroke by Presence of Breast Arterial Calcification. Risk for (**A**) acute myocardial infarction, (**B**) heart failure, and (**C**) stroke each significantly varied by the presence of breast arterial calcification (p<0.001) when comparing the unadjusted Kaplan-Meier Curves. BAC indicates breast arterial calcification; BAC+, presence of BAC and BAC-, absence of BAC.

**Supplementary Table 1.**
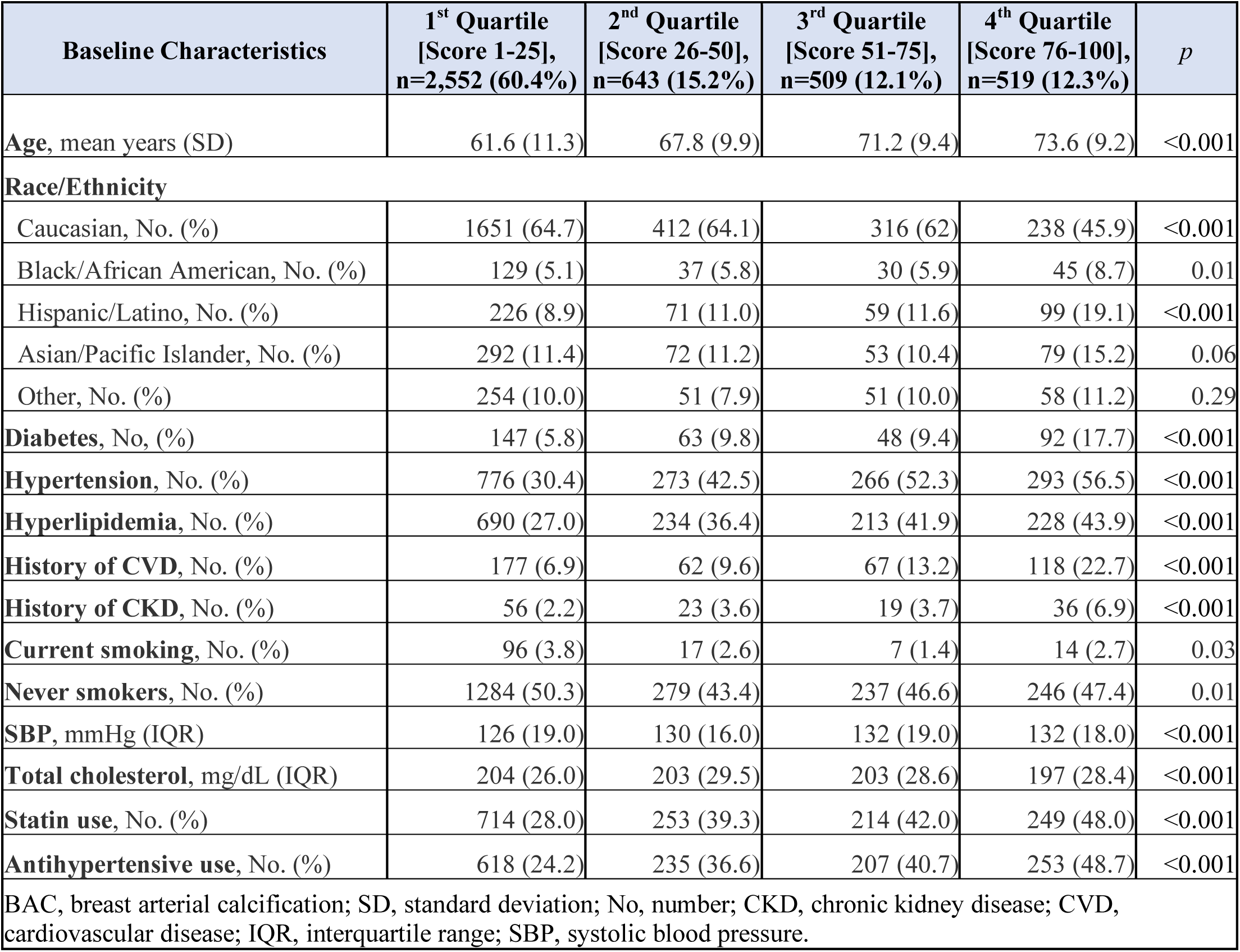
Baseline Participant Characteristics by Bradley Score Quartiles.

**Supplementary Table 2.** Association of Breast Arterial Calcification Presence and Cardiovascular Outcomes.

|  | Heart Failure<br>(n=298/17,911)<br>HR (95% CI) | *p* | Myocardial<br>Infarction<br>(n=83/18,051)<br>HR (95% CI) | *p* | Stroke<br>(n=259/17,914)<br>HR (95% CI) | *p* |
| --- | --- | --- | --- | --- | --- | --- |
| **Among all participants** |  |  |  |  |  |  |
| Model 1 | **1.51 (1.33, 1.73)** | **<0.001** | 1.11 (0.96, 1.28) | 0.184 | **1.29 (1.13, 1.48)** | **<0.001** |
| Model 2 | **1.46 (1.28, 1.66)** | **<0.001** | 1.10 (0.95, 1.28) | 0.200 | **1.25 (1.09, 1.43)** | **0.001** |
| Model 3 | **1.41 (1.24, 1.61)** | **<0.001** | 1.10 (0.95, 1.27) | 0.214 | **1.22 (1.07, 1.40)** | **0.003** |

Model 1: unadjusted. Model 2: adjusted for age and race/ethnicity. Model 3: adjusted for age, race/ethnicity, systolic blood pressure, diastolic blood pressure, diabetes, total cholesterol, low-density lipoprotein cholesterol, history of cardiovascular disease, history of chronic kidney disease, and smoking status. HR, hazard ratio; CI, confidence interval.

**Supplementary Table 3.** Association of The Bradley Score Quartiles and Cardiovascular Outcomes.

|  | Heart Failure<br>(n=298/17,911)<br>HR (95% CI) | *p* | Myocardial<br>Infarction<br>(n=83/18,051)<br>HR (95% CI) | *p* | Stroke<br>(n=259/17,914)<br>HR (95% CI) | *p* |
| --- | --- | --- | --- | --- | --- | --- |
| BAC negative, n=13,869 | *Referent* | -- | *Referent* | -- | *Referent* | -- |
| 1<sup>st</sup> Quartile [Score 1-25], n=2552 | **1.17 (1.00, 1.38)** | **0.049** | 1.03 (0.86, 1.23) | 0.767 | 1.06 (0.90, 1.25) | 0.501 |
| 2<sup>nd</sup> Quartile [Score 26-50], n=643 | **1.38 (1.03, 1.85)** | **0.029** | 1.23 (0.88, 1.71) | 0.232 | **1.35 (1.00, 1.82)** | **0.048** |
| 3<sup>rd</sup> Quartile [Score 51-75], n=509 | **2.03 (1.49, 2.76)** | **<0.001** | 1.14 (0.79, 1.66) | 0.485 | 1.19 (0.85, 1.67) | 0.322 |
| 4<sup>th</sup> Quartile [Score 76-100], n=519 | **1.70 (1.23, 2.35)** | **0.001** | 1.12 (0.77, 1.63) | 0.563 | **1.76 (1.27, 2.44)** | **<0.001** |

All data from the multivariable-adjusted model (Model 3), which adjusted for age, race/ethnicity, systolic blood pressure, diastolic blood pressure, diabetes, total cholesterol, low-density lipoprotein cholesterol, history of cardiovascular disease, history of chronic kidney disease, and smoking status. HR, hazard ratio; CI, confidence interval.

